# Frontline healthcare workers’ knowledge and perception of COVID-19 and willingness to work during the pandemic in Nepal: a nationwide cross-sectional web-based study

**DOI:** 10.1101/2020.08.12.20173609

**Authors:** Dipak Prasad Upadhyaya, Rajan Paudel, Daniel J. Bromberg, Dilaram Acharya, Kaveh Khoshnood, Kwan Lee, Ji-Huyuk Park, Seok-Ju Yoo, Archana Shrestha, BC Bom, Sabin Bhandari, Ramgyan Yadav, Ashish Timalsina, Chetan Nidhi Wagle, Brij Kumar Das, Ramesh Kunwar, Binaya Chalise, Deepak Raj Bhatta, Mukesh Adhikari

**Affiliations:** School of Medicine, Case Western Reserve University, Euclid Ave, Cleveland USA; Central Department of Public Health, Institute of Medicine, Tribhuvan University, Kathmandu, Nepal; Department of Social and Behavioral Sciences, Yale School of Public Health, Yale University, New Haven, CT, USA; Yale Center for Interdisciplinary Research on AIDS, Yale School of Public Health, New Haven, CT, USA; Department of Preventive Medicine, College of Medicine, Dongguk University, South Korea; Department of Epidemiology of Microbial Diseases, Yale School of Public Health, Yale University, New Haven CT, USA; Department of Public Health, Kathmandu University, Kathmandu, Nepal; Ministry of Health and Population, Kathmandu, Nepal; B.P Koirala Institute of Health Sciences, Dharan, Nepal; Hiroshima University, Graduate School for International Development and Cooperation, Hiroshima, Japan; Department of Health Policy and Management, Yale School of Public Health, Yale University, New Haven CT, USA

**Keywords:** COVID-19, Health-workers, Knowledge, Perception, Willingness

## Abstract

**Background:** The health sector’s effectiveness during a pandemic primarily depends on the availability, knowledge, skills, perceptions, and motivations of frontline healthcare workers. In this study, we aimed to investigate the contextual factors associated with the knowledge, perceptions, and the willingness of frontline healthcare workers to work during the COVID-19 pandemic in Nepal.

**Methods:** A total of 1051 frontline health-workers from all seven Nepalese provinces were included in this web-based cross-sectional study, which was conducted in May 2020. Using a 5-point Likert scale questionnaire, we collected information on knowledge, perceptions, and the willingness of frontline healthcare workers to work during the COVID-19 pandemic. Multivariable logistic regression was applied to identify independent associations between predictors and outcome variables.

**Results:** Of the 1051 frontline health-workers, 17.2% were found to have inadequate knowledge on COVID-19, 63.6% reported unsatisfactory perceptions of government response, and 35.9% showed an unwillingness to work during the pandemic. Health workers at local health facilities (AOR: 0.35; 95% CI: 0.17-0.68) and those with chronic diseases were less likely to have adequate knowledge of COVID-19. Nurses (AOR: 2.10; 95% CI: 1.38-3.18), health-workers from Karnali Province (AOR: 2.62; 95% CI: 1.52-4.53), and those who had adequate knowledge of COVID-19 (AOR: 3.86; 95% CI: 2.51-6.16) were more likely to have satisfactory perception towards government response to COVID-19. In addition, laboratory-workers, health workers from Karnali province, and those with adequate knowledge (AOR: 1.81; 95% CI: 1.27-2.58) were more likely to work during the COVID-19 pandemic.

**Conclusions:** We concluded that frontline healthcare workers have some gaps in knowledge-related to COVID-19; about two-thirds of them had a negative perception of government response, and nearly one-third of them were unwilling to work. These observations demonstrate that prompt actions are required to improve health-worker knowledge of COVID-19, address negative perceptions to government responses, and motivate them to provide healthcare services during the pandemic.

## Introduction

In December 2019, an outbreak of a pneumonia-like illness was first detected in Wuhan, Hubei Province of China[1], and subsequently, faced with an escalating number beyond China, the World Health Organization (WHO) declared the outbreak a pandemic[2]. Based on available evidence, the disease is transmitted between individuals via nasopharyngeal droplets or saliva. Furthermore, no vaccine or effective treatment for COVID-19 is currently available[3]. Nepal is a small country in South Asia that shares a border with China and observed its first case of COVID-19 on January 25, 2020 [4]. In the first half of May 2020, Nepal experienced an explosive increase in cases; more than three-fourths of all cases recorded do date occurred during this period. As of May 24, 2020, Nepal reported 603 cases and three fatalities [5]. To tackle the COVID-19 pandemic, Nepal first sealed its border with China, and then suspended all international flights and, on March 23, implemented a country-wide comprehensive lock-down. On April 3, after encountering its first case of local transmission, Nepal began to utilize its resources more systematically [6].

Concern has been expressed that health systems in low-income countries like Nepal are not sufficiently resilient to tackle a crisis like that presented by COVID-19. Due to resource constraints and a weak health system structure, rapid diagnosis of suspected cases and contact tracing are challenging[6]. Studies have shown that knowledge of infectious diseases is greatest among doctors and nurses[7,8]. In addition, age, sex, educational status, and preexisting medical conditions have been shown to affect health worker (HW) knowledge of Middle East Respiratory Syndrome (MERS), and Severe Acute Respiratory Syndrome (SARS)[9,10]. The primary sources of information about COVID 19 are international health organizations such as the Center for Disease Control (CDC), WHO and Ministry of Health, and social media. Moreover, the effectiveness of healthcare sectors during public health emergencies primarily depends on the availability, motivation and skills of frontline healthcare workers, and thus knowledge, their perceived willingness to work during uncertain times is essential [11], because appropriate perceptions and willingness to work during a pandemic are prerequisites of HW motivation to provide necessary treatment and to take the preventive actions required to reduce pandemic’s impact. Studies have shown that factors such are perceived personal risks, availability of personal protective equipment, family care obligations, HW gender, type of employment, personal confidence, defined role, dissemination of timely information, appropriate training, and personal health problems, influence perceptions and willingness to work during pandemics [11–14]. In the present study, frontline Healthcare Workers were defined as doctors, nurses, paramedics, laboratory workers, pharmacists, pathologist, technical personnel, public health workers, and others directly involved in COVID-19 prevention and treatment that have direct contact with confirmed or suspected cases during patient intake, screening, inspection, testing, transport, treatment, nursing, specimen collection, or pathogen detection.

To provide healthcare services effectively, it is essential to assess and update HW’ knowledge and improve motivation, and willingness, which depends on various factors at the individual, and to system levels. The present study describes the actual HW scenarios factors associated with their knowledge of COVID-19, their reactions to government interventions, and, most importantly, their perceived willingness to work during the pandemic. This study also provides valuable and actionable information to Nepal’s policymakers to allow the judicious allocation of scarce resources in the short run. In the long term, this study guides for those developing policies and programs. That might be instrumental in ensuring preparedness to meet the challenges posed by similar crises. Given this background, we aimed to investigate the contextual factors associated with the perceptions and willingness to work among Nepalese front-line healthcare workers during the COVID-19 pandemic to improve the prevention and management of future similar outbreaks.

## Methods

### Study participants and Sampling

We conducted a cross-sectional study using an online questionnaire from May 2020, among HWs in Nepal in accord with the Checklist for Reporting Results of Internet Surveys (CHERRIES) [15]. All participating HWs were aged 18 to 60 years old and ranged from high-level officials of the Ministry of Health and Population to paramedics working at the grassroots level in all seven provinces on Nepal. The research questionnaire was distributed to HWs using the health workers’ network. As an initial step, we first appointed a doctor or public health professional in each of the seven provinces to act as a coordinator and co-investigator in the team. These seven coordinators then sent HWs known to them a link to our questionnaire and asked that these individuals send a Google link to other HWs they knew. Fischer’s arctanh transformation as a power of 90% and a minimum correlation of 0.1[16], showed that the minimum sample size required was n=1046.

### Survey Instrument and data collection

The online questionnaire included 33 questions on socioeconomic characteristics, HWs’ knowledge of COVID-19, perception toward government response toCOVID-19, and perceived willingness to work during the pandemic. The Responses were rated using a 5-point Likert Scale (“Strongly Agree,” “Agree,” “Neutral,” “Disagree” to “Strongly Disagree”).

The socio-demographic characteristics investigated included age, gender, ethnicity, and marital status. This section of the questionnaire also included questions about chronic diseases of HWs, their caretaking responsibilities for dependent family members, nature of the employment, and type of health facility at which they worked. Knowledge of COVID-19 was rated as “adequate” and “inadequate,” perception of government response as “satisfactory” and “unsatisfactory” and willingness to work as “willing” and “unwilling.” Knowledge of COVID-19 was assessed based on knowledge of the causative agent, mode of transmission, proper use of PPE, infection prevention measures, and public health impact of the pandemic. Reaction to government response was determined by assessing response effectiveness, timeliness of information provided, provision of supplies, support received from administrative staff, and elected representatives. Factors influencing willingness to work during the pandemic were risk of self infection, healthcare service rationing, the requirement to work overtime, working with untrained HWs, deployment to another duty station, family risk, and ability to choose whether to work or not during the pandemic.

The questionnaire was prepared based on national COVID-19 guidelines issued by the Ministry of Health and Population of Nepal[17] and World Health Organization resource center guidelines for HWs on COVID-19 [18]. A team of medical doctors, public health workers, and an academic assessed the questionnaire for validity and relevance. Before conducting the survey, we conducted a pilot study on 30 participants to assess the reliability of the questionnaire items. The analysis revealed an overall Cronbach’s alpha score of 0.77, indicating higher internal consistency[19]. The questionnaire was prepared as a Google Form, and Facebook Messenger was used to sending the Google form link to participants [20,21]. The questionnaire took approximately 10 minutes to complete. To maintain data confidentiality, only two research team members had access to the data repository, stored on a password-protected computer.

### Data Management and Statistical Analysis

The data collected was downloaded in the form of a spreadsheet and checked for duplications and technical errors. After confirming the completeness, we exported the data to R Studio Software for full analysis [22]. Socio-demographic characteristics were subjected to descriptive analysis using the table 1 package in R software, and results are presented as frequencies, percentages, or as means and standard deviations [23].

Univariate logistic regression analysis was used to assess factors associated with adequate knowledge, satisfaction with the government response, and willingness to work using the *finalfit* package in R [24]. Parsimonious multivariate models were created for each dependent variable and included independent variables found to be significant (p-value <0.05) by univariate analysis. Coefficients in the regression models were transformed into odds ratios with 95 % confidence intervals. P-values of <0.05 were considered significant.

### Ethics statement

Ethical approval for the study was obtained from the Nepal Health Research Council (approval no: 329/2020 P). The first page of the questionnaire detailed the study objective, benefits, and harm. HWs provided e-consent prior to participating in the study. Participants were informed that they could leave the study at any time. Participation was voluntary and anonymous.

## Results

### Socio-demographic characteristics

A total of 1051 HWs participated in the study, 725 (68%) men and 326 (31%) women. The response rate was 79%. Table 1 shows the socio-demographic characteristics of health care workers who participated in the study. Nearly 49% of the participants were aged between 20 to 30 years. More than half (57.4%) of the HWs were Brahmin or Chhetri. The majority of the participants were doctors (35.3 %) and nurses or midwives (16.5%). Highest percentage response was from Bagmati Province (19.4%), which contains the capital city Kathmandu. Detailed information about the provinces of Nepal is explained in Additional File. Nearly 60% of HWs were permanent employees. More than 25% worked in local-level public health facilities, such as health posts, primary health care centers, community health units, and urban health centers. More than 50% worked in hospitals, public hospitals (22.8%) followed by teaching hospitals and private hospitals. Nearly 20% of respondents used a health-related managerial agency at the federal, provincial, or local levels. 13.5% of HWs reported having a chronic disease; the most common of which were diabetes, heart disease, and chronic respiratory disease and 64% of HWs had family members of less than five years or more than 60 years who needed their care and support.

**Table 1.**
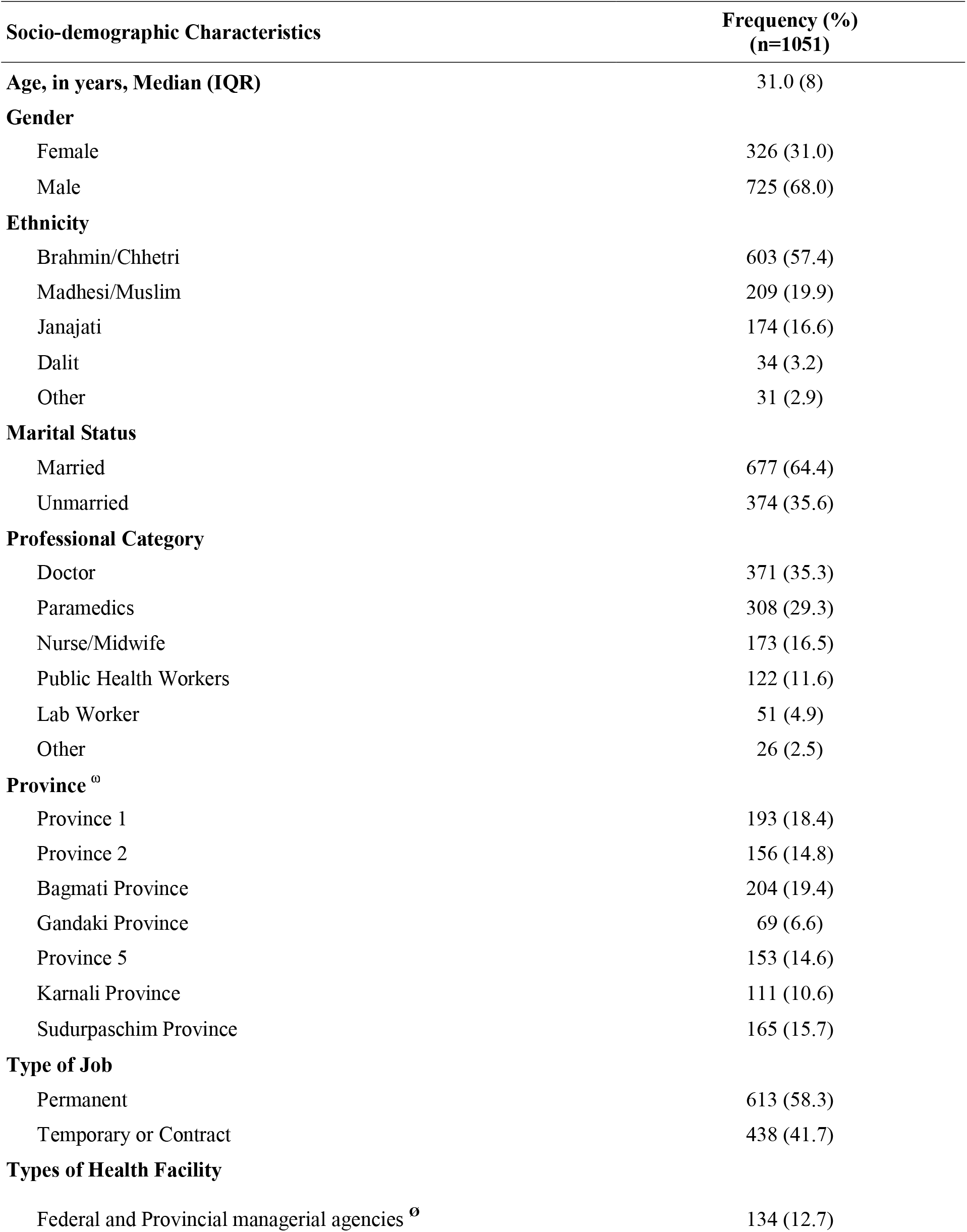
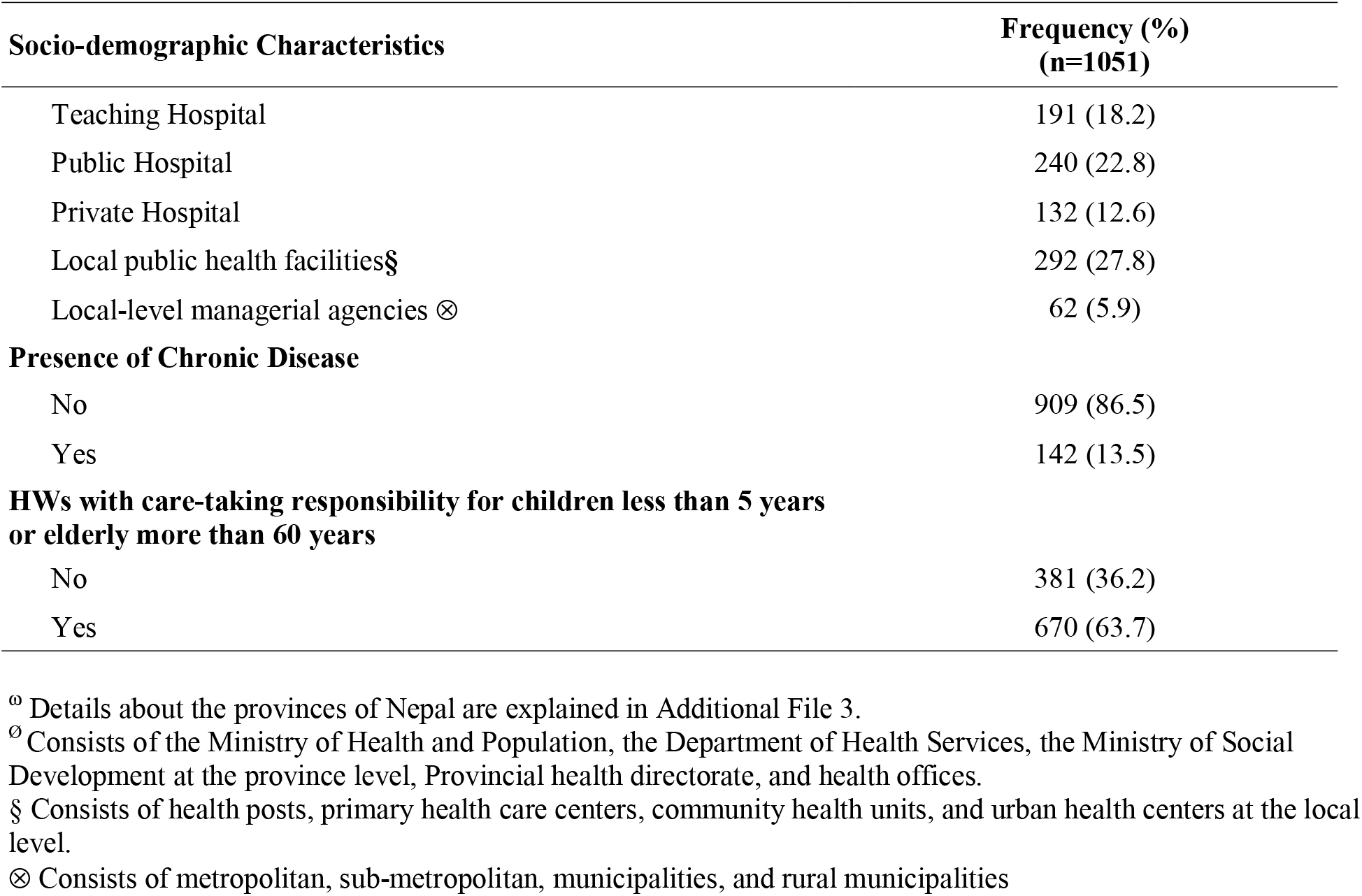
Sociodemographic Characteristics of Health workers in Nepal.

| Socio-demographic Characteristics | Frequency (%)<br>(n=1051) |
| --- | --- |
| **Age, in years, Median (IQR)** | 31.0 (8) |
| **Gender** |  |
| Female | 326 (31.0) |
| Male | 725 (68.0) |
| **Ethnicity** |  |
| Brahmin/Chhetri | 603 (57.4) |
| Madhesi/Muslim | 209 (19.9) |
| Janajati | 174 (16.6) |
| Dalit | 34 (3.2) |
| Other | 31 (2.9) |
| **Marital Status** |  |
| Married | 677 (64.4) |
| Unmarried | 374 (35.6) |
| **Professional Category** |  |
| Doctor | 371 (35.3) |
| Paramedics | 308 (29.3) |
| Nurse/Midwife | 173 (16.5) |
| Public Health Workers | 122 (11.6) |
| Lab Worker | 51 (4.9) |
| Other | 26 (2.5) |
| **Province <sup>o</sup>** |  |
| Province 1 | 193 (18.4) |
| Province 2 | 156 (14.8) |
| Bagmati Province | 204 (19.4) |
| Gandaki Province | 69 (6.6) |
| Province 5 | 153 (14.6) |
| Karnali Province | 111 (10.6) |
| Sudurpaschim Province | 165 (15.7) |
| **Type of Job** |  |
| Permanent | 613 (58.3) |
| Temporary or Contract | 438 (41.7) |
| **Types of Health Facility** |  |
| Federal and Provincial managerial agencies <sup>o</sup> | 134 (12.7) |

| **Socio-demographic Characteristics** | **Frequency (%)**<br>**(n=1051)** |
| --- | --- |
| Teaching Hospital | 191 (18.2) |
| Public Hospital | 240 (22.8) |
| Private Hospital | 132 (12.6) |
| Local public health facilities§ | 292 (27.8) |
| Local-level managerial agencies ⊗ | 62 (5.9) |
| **Presence of Chronic Disease** |  |
| No | 909 (86.5) |
| Yes | 142 (13.5) |
| **HWs with care-taking responsibility for children less than 5 years**<br>**or elderly more than 60 years** |  |
| No | 381 (36.2) |
| Yes | 670 (63.7) |

<sup>ⓐ</sup> Details about the provinces of Nepal are explained in Additional File 3.
<sup>ⓑ</sup> Consists of the Ministry of Health and Population, the Department of Health Services, the Ministry of Social
Development at the province level, Provincial health directorate, and health offices.
<sup>§</sup> Consists of health posts, primary health care centers, community health units, and urban health centers at the local
level.
<sup>⊗</sup> Consists of metropolitan, sub-metropolitan, municipalities, and rural municipalities

### Health Worker’ Knowledge of COVID-19

More than 80% of HWs had adequate knowledge of COVID-19 (See Table 2), and the percentage of men with adequate knowledge was higher than that of women. No significant difference in knowledge was observed among ethnic groups. However, significant differences were observed among health professionals. More than 90% of public health workers had adequate knowledge, while only 61.5% of other health workers such as Ayurveda—an ancient medical system prevalent in Nepal [25] and pharmacists had adequate knowledge. No provincial differences in COVID-19 knowledge were observed. However, knowledge of COVID-10 differed among HWs employed at different health facility types. No difference in COVID-19 related knowledge was found between those with or without chronic diseases or caretaking responsibilities. Multivariate logistic regression analysis showed that gender, professional category, and type of healthcare facility were associated with adequate knowledge of COVID-19, as shown in Table 2. Males were more likely to have adequate knowledge (OR: 1.60; 95 % CI: 1.02-2.47) than females. HWs in “other” professional categories such as pharmacists, and Ayurveda—had less adequate knowledge than doctors (OR: 0.33; 95 % CI: 0.14-0.80). HWs working at local health facilities were more likely to have inadequate knowledge about COVID-19 than those working at federal or provincial agencies (OR: 0.35; 95 % CI 0.17-0.68).

**Table 2.**
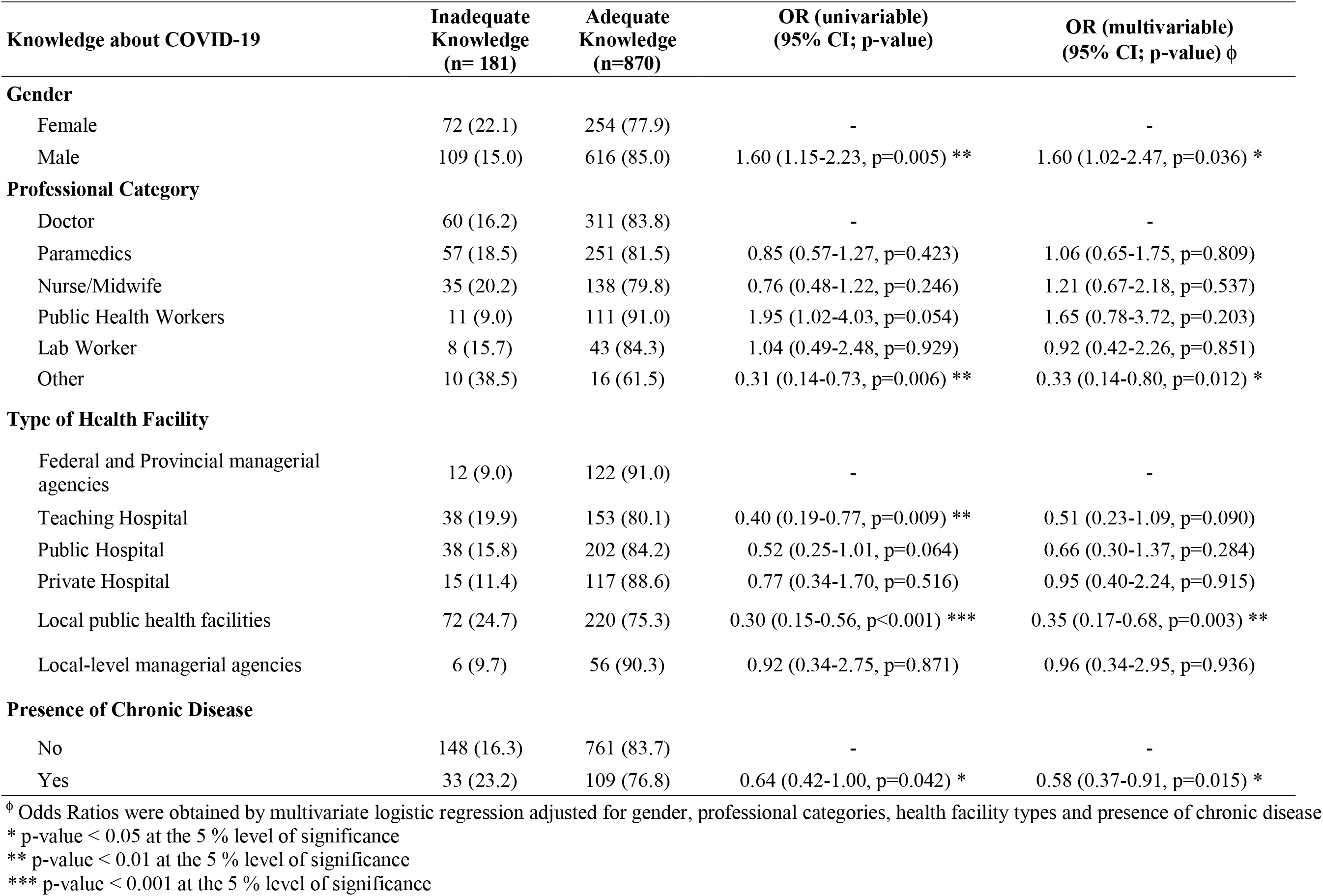
Factors Associated with Knowledge of COVID-19.

### Health Workers’ reactions to government response to COVID-19 Pandemic

More than 60% of HWs considered government response to COVID-19 Pandemic was unsatisfactory (Table 3). Gender and ethnicity were not found to influence perceptions of government response significantly. Nearly 74% of doctors reported government response to be unsatisfactory, while only 43 % of public health workers thought so. About 72 % of HWs from Bagmati Province and Province 2 were dissatisfied with the government response. Chronic disease and caretaking responsibility did not influence reactions to government response.

Multivariate logistic regression showed the reactions of HWs to government response were associated with a professional category, province, type of health facility, and adequacy of knowledge about COVID-19 (Table 3). Nurses were more likely to consider government response satisfactory than doctors (OR: 2.10; 95 % CI: 1.38-3.18). Similarly, public health professionals were more likely to consider government response to COVID-19 was satisfactory than doctors (OR: 1.83; CI 1.07 – 3.11). HWs from Province 6 (OR:2.62; 95 % CI: 1.52-4.53) and Province 7 (OR: 1.72; CI: 1.06-2.80) were more likely to consider government response satisfactory than those from Bagmati Province. Those working in public and teaching hospitals and local public health facilities were less likely to consider government response satisfactory than HW working from federal and provincial-level agencies. Interestingly, HWs with adequate knowledge of COVID-19 were more likely to consider government response satisfactory (OR: 3.86; 95 % CI 2.51-6.16).

**Table 3:** Factors Associated with Self-reported Perception of Government Response to COVID-19 Pandemic.

| Self-reported perception of government response ‡ | Unsatisfactory Government response (n=668) | Satisfactory Government response (n=383) | OR (univariable) | OR (multivariable) <sup>ϕ</sup> |
| --- | --- | --- | --- | --- |
| **Ethnicity** |  |  |  |  |
| Brahmin/Chhetri | 367 (60.9) | 236 (39.1) | - | - |
| Madhesi/Muslim | 145 (69.4) | 64 (30.6) | 0.69 (0.49-0.96, p=0.028) * | 1.15 (0.70-1.89, p=0.586) |
| Janajati | 115 (66.1) | 59 (33.9) | 0.80 (0.56-1.13, p=0.211) | 0.96 (0.65-1.42, p=0.846) |
| Dalit | 24 (70.6) | 10 (29.4) | 0.65 (0.29-1.34, p=0.260) | 0.67 (0.29-1.46, p=0.332) |
| Other | 17 (54.8) | 14 (45.2) | 1.28 (0.61-2.64, p=0.504) | 1.13 (0.50-2.49, p=0.770) |
| **Professional Category** |  |  |  |  |
| Doctor | 274 (73.9) | 97 (26.1) | - | - |
| Paramedics | 198 (64.3) | 110 (35.7) | 1.57 (1.13-2.18, p=0.007) | 1.18 (0.78-1.79, p=0.439) |
| Nurse/Midwife | 96 (55.5) | 77 (44.5) | 2.27 (1.55-3.31, p<0.001) *** | 2.10 (1.38-3.18, p<0.001) *** |
| Public Health Workers | 52 (42.6) | 70 (57.4) | 3.80 (2.49-5.85, p<0.001) *** | 1.83 (1.07-3.11, p=0.027) * |
| Lab Worker | 30 (58.8) | 21 (41.2) | 1.98 (1.07-3.60, p=0.027) | 1.52 (0.79-2.90, p=0.207) |
| Other | 18 (69.2) | 8 (30.8) | 1.26 (0.50-2.89, p=0.606) | 1.37 (0.52-3.38, p=0.506) |
| **Province** |  |  |  |  |
| Bagmati Province | 147 (72.1) | 57 (27.9) | - | - |
| Province 1 | 136 (70.5) | 57 (29.5) | 1.08 (0.70-1.67, p=0.726) | 0.99 (0.62-1.59, p=0.976) |
| Province 2 | 113 (72.4) | 43 (27.6) | 0.98 (0.61-1.56, p=0.937) | 0.88 (0.48-1.61, p=0.680) |
| Gandaki Province | 42 (60.9) | 27 (39.1) | 1.66 (0.93-2.93, p=0.083) | 1.69 (0.92-3.11, p=0.090) |
| Province 5 | 96 (62.7) | 57 (37.3) | 1.53 (0.98-2.40, p=0.062) | 1.48 (0.92-2.40, p=0.105) |
| Karnali Province | 46 (41.4) | 65 (58.6) | 3.64 (2.25-5.96, p<0.001) *** | 2.62 (1.52-4.53, p=0.001) ** |
| Sudurpaschim Province | 88 (53.3) | 77 (46.7) | 2.26 (1.47-3.49, p<0.001) *** | 1.72 (1.06-2.80, p=0.030) * |
| **Type of Health Facility** |  |  |  |  |
| Federal and Provincial managerial agencies | 53 (39.6) | 81 (60.4) | - | - |
| Teaching Hospitals | 135 (70.7) | 56 (29.3) | 0.27 (0.17-0.43, p<0.001) *** | 0.52 (0.29-0.93, p=0.027) * |
| Public Hospitals | 168 (70.0) | 72 (30.0) | 0.28 (0.18-0.43, p<0.001) *** | 0.41 (0.24-0.70, p=0.001) ** |
| Private Hospitals | 90 (68.2) | 42 (31.8) | 0.31 (0.18-0.50, p<0.001) *** | 0.52 (0.28-0.94, p=0.032) |
| Local public health facilities | 196 (67.1) | 96 (32.9) | 0.32 (0.21-0.49, p<0.001) *** | 0.49 (0.30-0.81, p=0.005) ** |
| Local-level managerial agencies | 26 (41.9) | 36 (58.1) | 0.91 (0.49-1.68, p=0.752) | 1.12 (0.58-2.20, p=0.742) |
| **Knowledge about COVID-19 †** |  |  |  |  |
| Inadequate | 154 (85.1) | 27 (14.9) | - | - |
| Adequate | 514 (59.1) | 356 (40.9) | 3.95 (2.61-6.20, p<0.001) *** | 3.86 (2.51-6.16, p<0.001) *** |

<sup>ϕ</sup> Odds Ratios were obtained by multivariate logistic regression adjusted for Ethnicity, Professional category, Province, type of health facility, and health worker perceived knowledge of COVID-19
† Knowledge about COVID-19 was self-reported; a combined score above average was regarded as “Adequate” knowledge and a score below or equal to the average was considered “Inadequate”.
‡ Self-reported perception of government response during COVID-19 pandemic was self-reported; a combined score above average was regarded as “Satisfactory perception” of knowledge and a score below or equal to the average was considered “Unsatisfactory perception”
\* p-value < 0.05 at the 5% level of significance
\*\* p-value < 0.01 at the 5% level of significance
\*\*\* p-value < 0.001 at the 5% level of significance

### Health workers’ willingness to work during the COVID-19 Pandemic

About 64 % of HWs reported a willingness to work under the challenging conditions created during the COVID-19 Pandemic (Table 4). No significant difference was observed between men and women with respect to willingness to work. About 74 % of laboratory workers were willing to work whereas, only 48.5 % of doctors were willing to do so. Furthermore, differences were observed between the seven provinces; ≃85 % of HWs in Karnali Province, but only 54.5 % of HWs from Province 2 were willing to work.

Multivariate analysis showed a willingness to work was associated with the professional category, province, presence of chronic disease, caregiving responsibility, and knowledge of COVID-19. Laboratory staffs (OR: 3.54; 95 % CI: 1.77-7.61), paramedics (OR: 2.52; 95 % CI: 1.79 – 3.58), public health workers (OR: 2.40; 95 % CI: 1.47-4.01), and nurses/midwives (OR: 2.09; 95 % CI: 1.40-3.47), were more willing to work during the pandemic than doctors. HWs from Karnali Province (OR: 2.96; 95 % CI: 1.62-5.64), and Sudurpaschim Province (OR: 2.10; 95 % CI: 1.28-3.48) were more likely to report willingness to work than those from Bagmati Province. HWs with responsibility for dependent family members were less willing to work than those without these responsibilities (OR: 0.72; 0.54-0.95). Finally, the HWs with adequate knowledge of COVID-19 were more prepared to work than those with inadequate knowledge (OR: 1.81; 1.27- 2.55).

**Table 4:** Factors Associated with Self-reported Willingness to Work during the COVID-19 Pandemic.

| Perceived Willingness to work | Unwilling to work<br>(n=377) | Willing to work<br>(n=674) | OR (univariable)<br>(95% CI, p-value) | OR (multivariable)<br>(95% CI, p-value) <sup>‡</sup> |
| --- | --- | --- | --- | --- |
| **Ethnicity** |  |  |  |  |
| Brahmin/Chhetri | 203 (33.7) | 400 (66.3) | - | - |
| Madhesi/Muslim | 89 (42.6) | 120 (57.4) | 0.68 (0.50-0.95, p=0.021) * | 1.11 (0.69-1.80, p=0.659) |
| Janajati | 69 (39.7) | 105 (60.3) | 0.77 (0.55-1.10, p=0.145) | 0.90 (0.62-1.31, p=0.575) |
| Dalit | 7 (20.6) | 27 (79.4) | 1.96 (0.88-4.95, p=0.121) | 1.74 (0.76-4.53, p=0.215) |
| Other | 9 (29.0) | 22 (71.0) | 1.24 (0.58-2.89, p=0.595) | 0.97 (0.43-2.36, p=0.945) |
| **Professional Category** |  |  |  |  |
| Doctor | 191 (51.5) | 180 (48.5) | - | - |
| Paramedics | 83 (26.9) | 225 (73.1) | 2.88 (2.09-3.99, p<0.001) *** | 2.52 (1.79-3.58, p<0.001) *** |
| Nurse/Midwife | 52 (30.1) | 121 (69.9) | 2.47 (1.69-3.64, p<0.001) *** | 2.09 (1.40-3.17, p<0.001) *** |
| Public Health Workers | 28 (23.0) | 94 (77.0) | 3.56 (2.26-5.77, p<0.001) *** | 2.40 (1.47-4.01, p=0.001) ** |
| Lab Worker | 11 (21.6) | 40 (78.4) | 3.86 (1.98-8.11, p<0.001) *** | 3.54 (1.77-7.61, p=0.001) ** |
| Other | 12 (46.2) | 14 (53.8) | 1.24 (0.56-2.79, p=0.600) | 1.24 (0.54-2.89, p=0.609) |
| **Province** |  |  |  |  |
| Bagmati Province | 90 (44.1) | 114 (55.9) | - | - |
| Province 1 | 72 (37.3) | 121 (62.7) | 1.33 (0.89-1.99, p=0.168) | 1.18 (0.77-1.81, p=0.446) |
| Province 2 | 71 (45.5) | 85 (54.5) | 0.95 (0.62-1.44, p=0.792) | 0.83 (0.47-1.45, p=0.510) |
| Gandaki Province | 29 (42.0) | 40 (58.0) | 1.09 (0.63-1.90, p=0.762) | 1.24 (0.69-2.22, p=0.473) |
| Province 5 | 64 (41.8) | 89 (58.2) | 1.10 (0.72-1.68, p=0.666) | 0.87 (0.56-1.37, p=0.554) |
| Karnali Province | 17 (15.3) | 94 (84.7) | 4.37 (2.48-8.06, p<0.001) *** | 2.96 (1.62-5.64, p=0.001) ** |
| Sudurpaschim Province | 34 (20.6) | 131 (79.4) | 3.04 (1.92-4.90, p<0.001) *** | 2.10 (1.28-3.48, p=0.004) ** |
| **Presence of Chronic Disease** |  |  |  |  |
| No | 313 (34.4) | 596 (65.6) | - | - |

| Perceived Willingness to work | Unwilling to work<br>(n=377) | Willing to work<br>(n=674) | OR (univariable)<br>(95% CI, p-value) | OR (multivariable)<br>(95% CI, p-value) <sup>♢</sup> |
| --- | --- | --- | --- | --- |
| Yes | 64 (45.1) | 78 (54.9) | 0.64 (0.45-0.92, p=0.015) * | 0.67 (0.46-0.99, p=0.043) * |
| **HWs having family members who need care** |  |  |  |  |
| No | 120 (31.5) | 261 (68.5) | - | - |
| Yes | 257 (38.4) | 413 (61.6) | 0.74 (0.57-0.96, p=0.026) * | 0.72 (0.54-0.95, p=0.021) * |
| **Perceived Knowledge about COVID-19<sup>†</sup>** |  |  |  |  |
| Inadequate | 86 (47.5) | 95 (52.5) | - | - |
| Adequate | 291 (33.4) | 579 (66.6) | 1.80 (1.30-2.49, p<0.001) *** | 1.81 (1.27-2.58, p=0.001) ** |
| **Perception of government response<sup>‡</sup>** |  |  |  |  |
| Unsatisfactory perception | 264 (39.5) | 404 (60.5) | - | - |
| Satisfactory Perception | 113 (29.5) | 270 (70.5) | 1.56 (1.20-2.05, p=0.001) ** | 1.12 (0.83-1.51, p=0.448) |

<sup>♢</sup> Odds Ratios were obtained by multivariate logistic regression adjusted for Ethnicity, Professional category, Province, Presence of Chronic Disease, health workers with family members requiring care, perceived knowledge of COVID-19, and perception of government response.
<sup>†</sup> Knowledge about COVID-19 was self-reported; a combined score above average was regarded as “Adequate” knowledge and a score below or equal to the average was considered “Inadequate”.
<sup>‡</sup> Perception of government response during COVID-19 pandemic was self-reported; a combined score above average was regarded as “Satisfactory perception” of knowledge and score below or equal to the average was considered “Unsatisfactory Perception”.
\* p-value < 0.05 at the 5 % level of significance
\*\* p-value < 0.01 at the 5 % level of significance
\*\*\* p-value < 0.001 at the 5 % level of significance

## Discussion

This is the first nationwide study on knowledge and perception of COVID-19 among frontline healthcare workers and their willingness to work during the pandemic in Nepal. About two in ten frontline healthcare workers (17.2%) had inadequate knowledge of COVID-19, which is higher than that reported in a Chinese study, in which ≃ 11 % demonstrated insufficient knowledge[26]. On the other hand, a study conducted by Bhagavathula S.A et al. reported that 61% of health workers had poor knowledge about COVID-19 transmission [27]. These differences between rates may have been due to variations in the level of knowledge accessed. Furthermore, the latter study was conducted in the first week of March 2020, and the Chinese study was conducted in the third week of May, when more information regarding COVID 19 was available and disseminated through different media. Knowledge is crucial for establishing perception and preventive behavior, which both affect coping interventions to some degree [28].

In addition, we found nearly two-thirds healthcare works (63.6%) believed government response to COVID-19 was unsatisfactory. A slightly higher level of satisfaction with government response was reported in a survey conducted on the Nepalese general public in April 2020 (71.4 %) [29]. The present stud also showed that most HWs (86%) experienced logistical shortcomings and reported inadequate supports form administrative (60%) and elected representatives (67.5%), which concurs with findings of a previous study[30]. Furthermore, our study shows that more than one in three HW (35.8%) were unwilling to work during the pandemic, which is considerable issues because the health system’s workload during the pandemic will be so high that all available health resources will be required to combat emergencies. In addition, the rate observed were higher than those reported in several other studies on willingness to work among health workers during public health emergencies[12–14,31,32]. In the present study, these high rates may have been due to inadequate knowledge (17%), preexisting chronic disease (13.5%), shortage of PPE (86%) and other factors [32,33]. The high rates of unwillingness to work during the pandemic revealed by our study demand the additional efforts be made to rectify the situation.

We also found that male health workers were more likely to report adequate knowledge; which is consistent with that found in another study conducted in Nepalese [34]. This finding may be due to greater interaction and socialization by men, and gendered norms, which means men are more likely to overestimate, and women are likely to underestimate personal knowledge [35–38]. The study also showed that pharmacists and Ayurveda had inadequate knowledge of COVID-19 rates as compared with doctors; finding is similar to a survey conducted in Nepal [34]. We also found that HWs in the local health facilities were less likely to have adequate knowledge than the HWs in federal or provincial agencies, which was possibly due to weaker implementation of COVID19 related governmental interventions at the local level than that at provincial or federal levels. In addition, HWs with a chronic disease considered they had inadequate knowledge of COVID19, perhaps because time limitations imposed by pre-existing conditions restricted studies about COVID-19.

This study shows that the professional category, province, type of health facility, and knowledge of COVID-19 were significantly associated with frontline health workers’ satisfaction with government response to the pandemic. Nurses were found to be more likely to be satisfied with government response than counterpart frontline doctors. This perception difference might have been due to the differences between levels of technical knowledge among doctors, nurses, and public health workers. Furthermore, health workers from Karnali and Sudurpaschim Provinces were more likely to be satisfied with government response than HWs from Bagmati province. However, the reasons responsible for these provincial variations were not determined. In addition, HWs from local public health facilities, teaching hospitals, and private hospitals had unsatisfactory perceptions than managerial level HWs at the ministry level, which we attribute to different work experiences, as HWs at health service outlets are directly exposed to risks and better understand the risks posed by logistical shortfalls than managerial level HWs. Interestingly, HWs with adequate knowledge of COVID-19 were more satisfied with government response than HWs with inadequate knowledge.

Interestingly, health workers professional category, province, presence of chronic disease, dependent family members, and knowledge about COVID 19 were associated with a willingness to work during the pandemic, and nurses, paramedics, public health workers, and laboratory staff were more willing to work than clinicians, which contradicts the results of a systematic review conducted by Aoyagi et al. [39]. HWs from Karnali and Sudurpaschim provinces were more willing to work than counterparts from Bagmati province. Similarly, it might be possible that due to virtually no cases of COVID-19 during the study, the HWs were willing to work in a humane way. Furthermore, HWs with adequate knowledge about COVID-19 were more willing to work, which concurs with a study performed on the 2007 influenza pandemic [40]. Our result shows that HWs with a chronic disease[41] and those that cared for family members[40] were less willing to work, which is also in line with previous studies [12,39]. It may be caring for family members and that coping with personal chronic health problems diminishes willingness to work [41].

This study was conducted to identify predictors of the willingness of frontline healthcare workers to work during the COVID-19 pandemic. The findings of this study can be used to inform various stakeholders and policymakers involved in the drafting of future interventions to improve the effectiveness of the health sector during public health crises. However, despite our efforts, this study has several limitations. First, data was obtained using a questionnaire over the web and health care workers were recruited using their personal networks. Therefore, our results should not be extended to healthcare workers that do not use the internet. Second, the data used was self-reported, which makes the study prone to desirability bias and inaccuracies. Furthermore, participants were asked to consider their willingness to work under hypothetical conditions that did not exist when Nepal comparatively observed a lower number of cases and fatalities. We recommend studies of the impacts of HW knowledge, perception, and willingness to work on health sector efficiency in the context of public health emergencies be undertaken.

## Conclusions

Health workers play an extremely critical role in the battle against pandemics. Therefore, their knowledge about the disease and their willingness to work are crucially required to prevent disease transmission and reduce morbidity and mortality. In view of the perceived knowledge gap of health workers about COVID-19, adequate training provides a means of addressing this shortcoming. The high-level dissatisfaction of HWs with logistical issues and shortages should also be at the focus of efforts to improving perceptions of government response. Health managers should be fully aware of the impacts of factors and devise comprehensive approaches that ensure the safety of HWs and promote coordination to motivate the HWs to work efficiently and effectively in a sustainable manner throughout the COVID-19 pandemic.

## Data Availability

It will be made available on request.

**List of Abbreviations**
COVID-19: Coronavirus Disease 2019
CI: Confidence Interval
HWs: Health Workers
MoHP: Ministry of Health and Population
OR: Odds Ratio
SARS-CoV-2: Severe Acute Respiratory Syndrome, Coronavirus 2
WHO: World Health Organization

## Declarations

### Ethics approval and consent to participate

The study protocol was approved by the Institutional Review Board of the Nepal Health Research Council (ERB Protocol Registration No. 329/2020 P). Electronic consent was required from all participants prior to completing the online questionnaire. A detailed description of the study was supplied on the first page of the online questionnaire.

### Consent for publication

Not applicable.

### Availability of data and materials

Data will be made available upon reasonable request by email to the corresponding author.

### Author Contributions

DPU, RP and MA conceptualized the study. DPU, RP, MA, AT, BC, and BB developed and pretested the questionnaire. BB, SB, RGY, RK, CNW, BKD, AT, MA, DRB, DPU, and RP supported data collection. DPU, MA, DJB, and RP performed the statistical analysis. DPU, MA and RP wrote the manuscript, and, DJB, KK, RP, KL, AS, JHP, SJY and DA edited, revised and finalized the manuscript. All authors contributed to the writing, editing, revision, and critical appraisal of the manuscript. All authors read and approved the final version of the manuscript.

## Acknowledgments

We authors thank Seema Subedi, a researcher at John Hopkins University, USA, for her suggestions during the conceptualization of the study. We wish to express our appreciation to all health workers that participated in the study. More importantly, we salute them for their tireless efforts to provide health services during the unprecedented challenge posed by COVID-19.

## Author Information

DPU is a graduate student at the School of Medicine, Case Western Reserve University, Euclid Ave, Cleveland USA, and faculty at Central Department of Public Health, Institute of Medicine, Tribhuvan University, Nepal. RP is an Assistant Professor at the Central Department of Public Health, Institute of Medicine, Tribhuvan University, Nepal. MA is an employee of the Ministry of Health and Population, and a graduate student at the Yale School of Public Health, Yale University, USA. DJB is a graduate student at the Yale School of Public Health, Yale University. KK is an associate professor at Yale School of Public Health, AS is an associate professor at the Department of Public Health, Kathmandu University, Nepal. AT, BB, RGY, CNW, BKD, RK, and DRB are employed by the Ministry of Health and Population, Government of Nepal. SB is an assistant professor at B.P Koirala Institute of Health Sciences, Dharan, Nepal. BC is a student at a Graduate School for International Development and Cooperation, Hiroshima University, Japan. KL, JHP, and SJY are professors at the Department of Preventive Medicine, College of Medicine, Dongguk University, Gyeongju, South Korea. DA is a recent doctoral graduate from the Department of Preventive Medicine, College of Medicine, Dongguk University, South Korea. All authors have read and understood the *ICMJE* criteria for authorship policy.

## References

1. Huang C, Wang Y, Li X, Ren L, Zhao J, Hu Y, et al. Clinical features of patients infected with 2019 novel coronavirus in Wuhan, China. The Lancet. 2020;395: 497–506. doi:10.1016/S0140-6736(20)30183-5

2. WHO Director-General’s opening remarks at the media briefing on COVID-19 - 11 March 2020. [cited 4 Jun 2020]. Available: https://www.who.int/dg/speeches/detail/who-directorgeneral-s-opening-remarks-at-the-media-briefing-on-covid-19---11-march-2020

3. Coronavirus. [cited 10 Jul 2020]. Available: https://www.who.int/westernpacific/healthtopics/coronavirus

4. Shrestha R, Shrestha S, Khanal P, Kc B. Nepal’s first case of COVID-19 and public health response. J Travel Med. 2020;27. doi:10.1093/jtm/taaa024

5. Corona Info- Ministry of Health and Population. [cited 10 Jul 2020]. Available: https://covid19.mohp.gov.np/#/

6. health-sector-emergency-response-plan-covid-19-endorsed-may-2020.pdf. Available: https://www.who.int/docs/default-source/nepal-documents/novel-coronavirus/health-sectoremergency-response-plan-covid-19-endorsed-may-2020.pdf?sfvrsn=ef831f44_2

7. Huynh G, Nguyen TNH, Tran VK, Vo KN, Vo VT, Pham LA. Knowledge and attitude toward COVID-19 among healthcare workers at District 2 Hospital, Ho Chi Minh City. Asian Pac J Trop Med. 2020;13: 260. doi:10.4103/1995-7645.280396

8. Olum R, Chekwech G, Wekha G, Nassozi DR, Bongomin F. Coronavirus Disease-2019: Knowledge, Attitude, and Practices of Health Care Workers at Makerere University Teaching Hospitals, Uganda. Front Public Health. 2020;8. doi:10.3389/fpubh.2020.00181

9. Khan MU, Shah S, Ahmad A, Fatokun O. Knowledge and attitude of healthcare workers about middle east respiratory syndrome in multispecialty hospitals of Qassim, Saudi Arabia. BMC Public Health. 2014;14: 1281. doi:10.1186/1471-2458-14-1281

10. Deng J-F, Olowokure B, Kaydos-Daniels SC, Chang H-J, Barwick RS, Lee M-L, et al. Severe acute respiratory syndrome (SARS): knowledge, attitudes, practices and sources of information among physicians answering a SARS fever hotline service. Public Health. 2006;120: 15–19. doi:10.1016/j.puhe.2005.10.001

11. Watt K, Tippett VC, Raven SG, Jamrozik K, Coory M, Archer F, et al. Attitudes to living and working in pandemic conditions among emergency prehospital medical care personnel. Prehospital Disaster Med. 2010;25: 13–19. doi:10.1017/s1049023x00007597

12. Martin SD. Nurses’ ability and willingness to work during pandemic flu. J Nurs Manag. 2011;19: 98–108. doi:10.1111/j.1365-2834.2010.01190.x

13. Hope K, Durrheim D, Barnett D, D’Este C, Kewley C, Dalton C, et al. Willingness of frontline health care workers to work during a public health emergency. Aust J Emerg Manag. 2010;25: 39–47.

14. Qureshi K, Gershon RRM, Sherman MF, Straub T, Gebbie E, McCollum M, et al. Health care workers’ ability and willingness to report to duty during catastrophic disasters. J Urban Health Bull N Y Acad Med. 2005;82: 378–388. doi:10.1093/jurban/jti086

15. Eysenbach G. Improving the Quality of Web Surveys: The Checklist for Reporting Results of Internet E-Surveys (CHERRIES). J Med Internet Res. 2004;6: e34. doi:10.2196/jmir.6.3.e34

16. Computational Statistics with R, Volume 32 - 1st Edition. [cited 10 Jul 2020]. Available: https://www.elsevier.com/books/computational-statistics-with-r/rao/978-0-444-63431-3

17. Coronavirus disease (COVID-19) outbreak updates & resource materials – Health Emergency Operation Center. [cited 10 Jul 2020]. Available: https://heoc.mohp.gov.np/update-on-novel-corona-virus-covid-19/

18. Technical guidance publications. [cited 10 Jul 2020]. Available: https://www.who.int/emergencies/diseases/novel-coronavirus-2019/technical-guidancepublications

19. Using and Interpreting Cronbach’s Alpha | University of Virginia Library Research Data Services + Sciences. [cited 10 Jul 2020]. Available: https://data.library.virginia.edu/using-and-interpreting-cronbachs-alpha/

20. Google Forms. [cited 10 Jul 2020]. Available: https://docs.google.com/forms/u/0/?usp=mkt_forms

21. Social Media Stats Nepal. In: StatCounter Global Stats [Internet]. [cited 10 Jul 2020]. Available: https://gs.statcounter.com/social-media-stats/all/nepal

22. R: The R Project for Statistical Computing. [cited 10 Jul 2020]. Available: https://www.r-project.org/

23. Using the table1 Package to Create HTML Tables of Descriptive Statistics. [cited 10 Jul 2020]. Available: https://cran.r-project.org/web/packages/table1/vignettes/table1-examples.html

24. Harrison E, Drake T, Ots R. finalfit: Quickly Create Elegant Regression Results Tables and Plots when Modelling. 2020. Available: https://CRAN.R-project.org/package=finalfit

25. Durkin M. Ayurvedic treatment for jaundice in Nepal. Soc Sci Med. 1988;27: 491–495. doi:10.1016/0277-9536(88)90372-3

26. Zhang M, Zhou M, Tang F, Wang Y, Nie H, Zhang L, et al. Knowledge, attitude, and practice regarding COVID-19 among healthcare workers in Henan, China. J Hosp Infect. 2020;105: 183–187. doi:10.1016/j.jhin.2020.04.012

27. Bhagavathula AS, Aldhaleei WA, Rahmani J, Mahabadi MA, Bandari DK. Knowledge and Perceptions of COVID-19 Among Health Care Workers: Cross-Sectional Study. JMIR Public Health Surveill. 2020;6: e19160. doi:10.2196/19160

28. McEachan R, Taylor N, Harrison R, Lawton R, Gardner P, Conner M. Meta-Analysis of the Reasoned Action Approach (RAA) to Understanding Health Behaviors. Ann Behav Med Publ Soc Behav Med. 2016;50: 592–612. doi:10.1007/s12160-016-9798-4

29. COVID 19, Citizen’s pulse (A National Survey on COVID 19-Nepal) – Participedia. [cited 10 Jul 2020]. Available: https://participedia.net/case/6543

30. Sim MR. The COVID-19 pandemic: major risks to healthcare and other workers on the front line. Occup Environ Med. 2020;77: 281–282. doi:10.1136/oemed-2020-106567

31. Kaiser HE, Barnett DJ, Hsu EB, Kirsch TD, James JJ, Subbarao I. Perspectives of future physicians on disaster medicine and public health preparedness: challenges of building a capable and sustainable auxiliary medical workforce. Disaster Med Public Health Prep. 2009;3: 210–216. doi:10.1097/DMP.0b013e3181aa242a

32. Balicer RD, Barnett DJ, Thompson CB, Hsu EB, Catlett CL, Watson CM, et al. Characterizing hospital workers’ willingness to report to duty in an influenza pandemic through threat-and efficacy-based assessment. BMC Public Health. 2010;10: 436. doi:10.1186/1471-2458-10-436

33. Yonge O, Rosychuk RJ, Bailey TM, Lake R, Marrie TJ. Willingness of university nursing students to volunteer during a pandemic. Public Health Nurs Boston Mass. 2010;27: 174–180. doi:10.1111/j.1525-1446.2010.00839.x

34. Knowledge, attitude and practice regarding COVID-19 among healthcare workers in Chitwan, Nepal. 2020 [cited 15 Jul 2020]. doi:10.21203/rs.3.rs-26774/v1

35. Cooper KM, Krieg A, Brownell SE. Who perceives they are smarter? Exploring the influence of student characteristics on student academic self-concept in physiology. Adv Physiol Educ. 2018;42: 200–208. doi:10.1152/advan.00085.2017

36. Ehrlinger J, Dunning D. How chronic self-views influence (and potentially mislead) estimates of performance. J Pers Soc Psychol. 2003;84: 5–17. doi:10.1037/0022-3514.84.1.5

37. Beyer S. Gender differences in the accuracy of self-evaluations of performance. J Pers Soc Psychol. 1990;59: 960–970. doi:10.1037/0022-3514.59.5.960

38. Female Medical Students Underestimate Their Abilities And Males Tend To Overestimate Theirs. In: ScienceDaily [Internet]. [cited 10 Jul 2020]. Available: https://www.sciencedaily.com/releases/2008/10/081003122713.htm

39. Aoyagi Y, Beck CR, Dingwall R, Nguyen-Van-Tam JS. Healthcare workers’ willingness to work during an influenza pandemic: a systematic review and meta-analysis. Influenza Other Respir Viruses. 2015;9: 120–130. doi:10.1111/irv.12310

40. Daugherty EL, Perl TM, Rubinson L, Bilderback A, Rand CS. Survey study of the knowledge, attitudes, and expected behaviors of critical care clinicians regarding an influenza pandemic. Infect Control Hosp Epidemiol. 2009;30: 1143–1149. doi:10.1086/648085

41. (PDF) Factors Related to Essential Workers’ Ability and Willingness to Work and Comply with Personal Infection Control Protocol During a Large Scale Influenza Pandemic in Hawaii. [cited 16 Jul 2020]. Available: https://www.researchgate.net/publication/256456823_Factors_Related_to_Essential_Workers'_Ability_and_Willingness_to_Work_and_Comply_with_Personal_Infection_Control_Protocol_During_a_Large_Scale_Influenza_Pandemic_in_Hawaii

